# Maternal Profiles Account for Birth Weight Differences Across Ethnicities: Results from Three Canadian Birth Cohorts

**DOI:** 10.1101/2025.06.02.25328798

**Authors:** Wei Q. Deng, Marie Pigeyre, Sandi M. Azab, Amel Lamri, Russell J de Souza, Natalie Williams, Kyla Belisario, Nathan Cawte, Dipika Desai, Katherine M. Morrison, Stephanie A. Atkinson, Koon K. Teo, Theo J. Moraes, Padmaja Subbarao, Stuart E. Turvey, Piush Mandhane, Elinor Simons, Guillaume Pare, Sonia S. Anand

## Abstract

**Background:** Marked differences in birth weight (BW) between South Asian and White European-origin populations are well-documented and pose public health concerns.

**Methods:** We analyzed fetal BW, the fat mass (FM), and fat-free mass (FFM) components in South Asian (*n*=938) and White European (*n*=3,044 and 804) newborns from three Canadian birth cohorts, examining the contribution of 16 maternal factors to observed BW differences using epidemiological and Mendelian randomization analyses.

**Findings:** South Asian newborns had on average, a significantly lower BW (3.3±0.4kg) than White Europeans (3.5±0.5kg), even after accounting for birth length (*p*<0.001). FFM was the primary driver of this difference, contributing to 0.22kg lower BW (*p*<2.2E-16), while FM had a significant but weaker counteracting effect of 0.01kg higher BW in South Asians (*p*=0.006). Five maternal factors demonstrated a direct maternal genetic influence: pre-pregnancy weight primarily increased BW via FFM, it also had a non-negligible increasing effect on FM. On the other hand, maternal glucose and gestational diabetes mellitus (GDM) causally increased BW through FM accumulation. Maternal height had a minimal effect only on FFM. After adjusting for these 5 maternal predictors, roughly 50% of the ethnic difference in BW (0.1kg; 95% CI: 0.067–0.13kg) was accounted for.

**Interpretation:** Different maternal factors influence specific components of BW. Targeting body fat reduction and maternal glucose regulation in South Asian mothers may help reduce the intergenerational transmission of increased FM and its associated adverse health outcomes.

**Funding:** This study was funded by the Canadian Institutes of Health Research DOHaD Team Grant: MWG-146332.

**Research in Context:** *Evidence before this study:* Ethnic difference in birth weight between South Asian (and other non-White European populations) and White European-origin populations are well-documented in cohort studies such as in Born in Bradford and nation-wide estimates, but not well understood. Previous studies have looked at different body compositions, maternal environment, or genetics, but no study has integrated all of these to determine the causal drivers of differences in BW, and their impact on body composition, between South Asians and White Europeans.

*Added value of this study:* The present study adds evidence that difference in birth weight is not a unitary concept but rather the balance of both fat and lean mass. Importantly, these components are strongly driven by a set of maternal factors, and accounting for these factors, we were able to level the difference in birth weight by up to 50%.

*Implications of all the available evidence:* Together with other emerging research on the causal determinants of birth weight, these findings suggest a potential set of maternal intrauterine factors that can be targeted to optimize early-life and long-term cardiometabolic health—particularly in high-risk populations such as South Asians. These results also emphasize the need to go beyond birth weight alone and consider neonatal body composition and its maternal drivers when evaluating and addressing health disparities.

## Introduction

Birth weight (BW) is a critical determinant of neonatal and long-term health^1–4^, with significant ethnic differences observed between South Asian and White European populations^5–7^. South Asian newborns tend to have lower BW and higher adiposity at birth compared to White Europeans, even after accounting for gestational age and maternal socioeconomic status^6,8^. The observed disparity emerged as a potential contributing factor to the markedly higher risk of cardiometabolic diseases observed in South Asian populations^9–11^. Indeed, emerging research leveraging genetics, causal inference, and longitudinal designs suggests that BW is not merely correlated with but can causally drive the risk for cardiovascular disease^2,12–15^ and type 2 diabetes (T2D)^16–20^. Thus, there is a pressing need to identify causal mechanisms driving the ethnic difference in BW and their health consequences.

South Asian newborns typically exhibit lower fat-free mass (FFM) and higher body fat percentage (BF%) than their White European counterparts^21^, a pattern that persists even after migration^22^. BF% can be robustly estimated using skinfold thickness^23^, which is characteristically different between White European and South Asian newborns, implicating maternal BF% and glucose concentration as potential risk factors^24^. This “lean-fat phenotype” suggests that BW differences may not solely reflect overall fetal growth but rather differences in body composition. The apparent association between a “lean-fat phenotype” and T2D susceptibility among South Asians^25,26^ also raises the question of whether a low BW or a high BF% equivalently contributes to the burden of cardiometabolic diseases in this population^1,9,27^.

Previous studies have looked at different body weight compartments (i.e. lean vs. fat mass)^21^, clinical risk factors^28–31^, maternal environment^32–39^, or genetics^29,40–43^, but no study has integrated all of these to determine the causal drivers of differences in BW between South Asians and White Europeans. Here we explore a three-step approach to this problem (1) characterization of the body compartments responsible for the difference in BW, (2) identification of maternal factors associated with these differences (i.e. at the compartment level) and (3) insights into potential causal pathways using principles of Mendelian randomization.

## Methods

### Study populations

Our analysis focused on three NutriGen birth cohorts with genome-wide genotype and cord blood DNA methylation data. The CHILD study is a prospective longitudinal birth cohort^44^ that enrolled pregnant women who gave birth between 2009 and 2012 across four Canadian cities, with participant recruitment designed to reflect the demographic diversity of the populations in those regions. The FAMILY study, based in Hamilton, Ontario, Canada, examined early atherosclerosis indicators in a predominantly White European population^45^. The START study exclusively enrolled mothers (2011–2013) who originated from the Indian subcontinents and resided in Ontario’s Peel region^46^. These cohorts were not enriched for clinical conditions. Each cohort provided a comprehensive suite of biological samples, clinical evaluations, and questionnaire-based data. Only singleton births were included in the present analysis. In the CHILD and FAMILY cohorts, which are predominantly composed of individuals of White European ancestry, we retained participants who self-identified as White European and also matching the inferred genetic ancestry. In the START cohort, we retained participants whose self-reported ethnicity and inferred genetic ancestry were both South Asian. Thereafter, all references to the CHILD and FAMILY cohorts refer specifically to the White European subsamples, and all references to the START cohort refer to the South Asian sample.

### Ethics

Ethical approval was obtained independently from the Hamilton Integrated Research Ethics Board (HiREB): CHILD (REB 07–2929) and START (REB 10–640). CHILD was additionally approved by the respective Human Research Ethics Boards at McMaster University, the Universities of Manitoba, Alberta, and British Columbia, and the Hospital for Sick Children. Legal guardians of each participant provided written informed consent. Written informed consent was obtained from the parent/guardian (participating mother) for each study separately. We also have now obtained additional ethics board approval from HiREB (REB 16592) for using the data from the two cohorts together without additional consent from the participants.

### Maternal measurements

We focused on 16 maternal and prenatal phenotypes from diet patterns, smoking exposure, demographics, adiposity and body size to other clinical variables. Three maternal diet patterns^47^ were identified using principal component analysis on food frequency questionnaires: i) plant-based: high in vegetables, legumes, and whole grains; ii) Western: high in sweets, red meats, and processed foods; and iii) health-conscious: rich in seafood, poultry, vegetables, and fruits. Maternal smoking history (current, quit during pregnancy, quit before pregnancy, never-smoker) and household smoking exposure (hours per week) were self-reported. Maternal education was harmonized across the cohorts and captured by the number of years in school. Socioeconomic variables were consolidated into a single continuous measure validated in the context of cardiometabolic disease risk^48^: the social disadvantage index (SDI), encompassing household income, marital status, and maternal employment. Maternal body size were measured by pre-pregnancy BMI (kg/m^2^), pre-pregnancy weight (kg), and height (cm). Clinical variables of interest included maternal age at delivery, gestational weight gain (kg), gestational diabetes mellitus (GDM) status, pre-eclampsia, and parity. In CHILD, GDM was defined by a combination of self-report of diagnosis at baseline or 1-year follow-up, or record of GDM on birth chart. In FAMILY, GDM was determined by self-report, diabetes therapies (insulin, oral glucose-lowering drugs, diabetic diet), birth chart records, and the IADPSG criteria^49^ (fasting glucose ≥5.1 mmol/L, 1-hour post-OGTT≥10.0 mmol/L, or 2-hour post-OGTT≥8.5 mmol/L). In START, GDM was additionally classified using Born in Bradford (BiB) criteria^50,51^ (OGTT glucose>=5.2 mmol/l at baseline or 2-hour post OGTT glucose>=7.2 mmol/l). In FAMILY and START, the area under the curve (AUC) is calculated from OGTT data using the trapezoidal rule to quantify the total increase in blood glucose levels throughout the test^52^ (glucose AUC). We used GDM status as a surrogate measure of glucose AUC in CHILD.

### Offspring characteristics

Gestational age at birth, reported in weeks and days, and newborn sex assigned at birth, were collected from participants’ birth charts. Offspring anthropometrics are available at birth from medical charts or measured at <2 days old, and subsequently measured at the 1, 2 (START only), 3, and 5-year follow-up visits. This included height (cm) and weight (g) at all visits; and the sum of skinfold thickness, previously derived from triceps and subscapular skinfolds (mm)^24,53^ at 3- and 5-year visits. Since gestational age at birth is strongly correlated with BW, only full-term newborns were retained (gestational age at birth greater or equal to 37 weeks). This yielded 3044, 803, and 938 mother-child dyads with complete gestational age and BW data.

### Estimating BF%, fat mass, and fat-free mass in newborns

Newborn BF% was estimated using Slaughter’s skinfold-thickness equations^53^ in FAMILY and START. Leveraging newborn metrics and maternal characteristics harmonized across the cohorts, we constructed a prediction model in FAMILY and applied the best model to estimate BF% in CHILD. Based on the predicted and estimated BF%, we calculated the fat mass (FM) by multiplying BW by BF%, and FFM by taking the difference between BW and FM. Details of the equations, model training, and validation can be found in Appendix A of **Supplementary Methods**.

### Genome-wide genotype data

Genome-wide genotype data and quality controls (QCs) steps have been described elsewhere^54,55^. In all cohorts, the genetic ancestry of participating mothers and children was ascertained by mapping genetic principal components (PCs) to global population references from the 1000 Genomes Project^56^. The genetic subset included 1451, 267, 533 mother-child dyads with complete gestational age and BW data in CHILD, FAMILY, and START, respectively (**Table S1**). Detailed data QCs are described in Appendix B of **Supplementary Methods**.

### Construction and validation of polygenic risk scores

We computed 10 fetal PGS (fPGS) for BW, height, FM, FFM, BMI, T2D, pre-eclampsia, fasting glucose level, education, and smoking, and maternal PGS of the same traits (mPGS) in all cohorts. These were chosen to mirror the 16 maternal and prenatal characteristics. All PGSs, except for GDM, were curated from the Polygenic Score Catalog^57^, selecting the most predictive PGSs for each target phenotype. All scores were standardized to have a mean of zero and a unit variance. Details on the score construction are available in Appendix B of **Supplementary Methods**.

### Statistical Analysis

We conducted statistical analyses in three stages. First, we established and confirmed difference in birth metrics (GA, birth length, BW, FM, FFM, ratios of BW components and birth length) across cohorts in the full sample with non-missing BW and GA data. A parametric analysis of variance (ANOVA) via an *F*-test was used to evaluate the significance of difference in birth outcomes. For subsequent analyses, we adjusted BW and its components by child’s sex, gestational age, and age of mother at delivery (except when examining age at delivery as a predictor). Second, we systematically screened each maternal predictor for its association with BW, FM, and FFM in univariate regression models. This step was carried out separately in each cohort and then meta-analyzed via a fixed effect model. To account for multiple hypothesis testing, *p*-values were adjusted using the Benjamini-Hochberg procedure^58^ to control for False Discovery Rate (FDR). Only maternal predictors associated with at least one birth component in the meta-analysis (FDR corrected *p*-value<0.05) with homogenous effect across cohorts were retained. To determine whether associated maternal factors causally influence BW, we leveraged genetic scores of these maternal factors as instrumental variables under a Mendelian Randomization framework. This approach reduces bias from confounding and reverse causation by using genetic proxies assigned at conception. Specifically, we conducted causal analyses using: 1) an inverse variance weighted (IVW) regression, and 2) the generalized methods of moments (GMM). We considered a maternal trait to direct influence BW if the genetically predicted maternal trait (i.e. mPGS) remained significantly associated with BW after adjusting for children genetics (i.e. fPGS). If the mPGS was not significantly associated with the mPGS target trait (e.g. mPGS of BMI should be strongly associated with maternal BMI), we selected an alternative mPGS that exhibited the strongest association with the target maternal trait to minimize weak instrument bias. Causal analysis was conducted separately in each cohort and then meta-analyzed using a fixed-effect model. We conducted additional sensitivity analysis by repeating the causal analysis in mother without GDM, using alternative definitions for GDM in FAMILY and START, and also explored the robustness of the findings by reversing the roles the role of fetal and maternal PGSs. After identifying the causal factors, we assessed whether accounting for these factors could explain the observed difference in BW using two complementary approaches: (1) an adjusted regression model that included these factors as covariates, and (2) a matched analysis, in which SA and White European mothers were matched on the identified factors and unmatched mothers were excluded. Matching was performed using the *MatchIt* R package with the optimal pair matching, based on four covariates (excluding maternal height to retain a reasonable number of paired samples), and exact matching on pre-pregnancy BMI and GDM status. In both cases, ANOVA was used to evaluate the significance of the remaining difference in birth outcomes. All data processing and analyses were conducted in R v.4.1.0 ^59^.

## Results

### Study sample characteristics

We noted significant between-cohort differences in BW, GA, and birth length (*p* < 0.001; **Table 1**). South Asian newborns had on average, a significantly lower BW (3.3±0.4kg) than White Europeans (3.5±0.5kg), which persisted after accounting for birth length (*p*<0.001; **Figure 1**). Further, South Asian newborns also had a shorter gestation at birth (39.4±1.1 weeks) than White European newborns in CHILD (39.7±1.1weeks; *p*<0.001) but not compared to those in FAMILY (39.4±1.2 weeks; **Table 1; Figure S1**).

**Figure 1.**
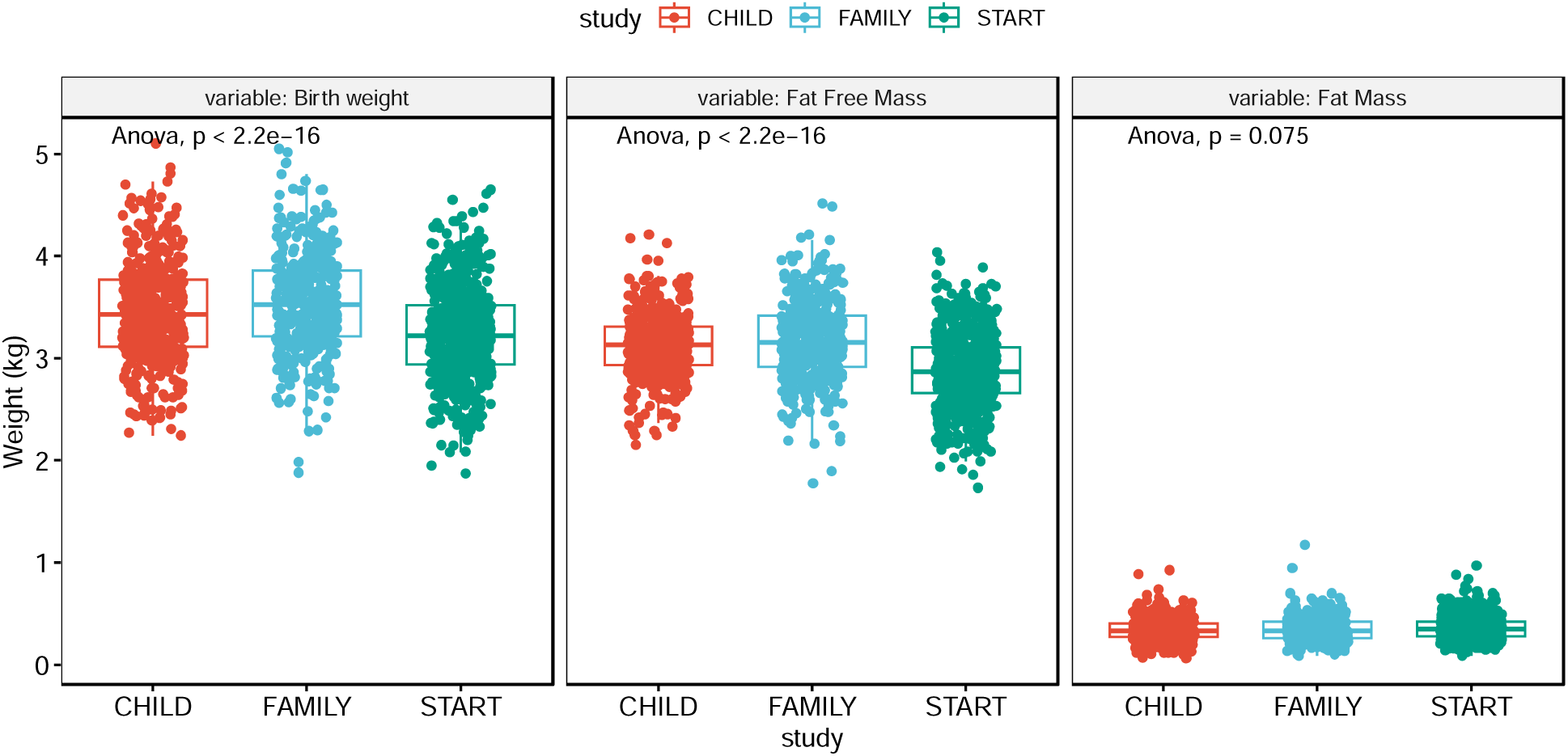
Stratified boxplots of birth weight, fat-free mass, and fat mass across three studies: CHILD, FAMILY, and START. Each boxplot illustrates the distribution of outcomes within each study cohort. Pairwise comparisons between studies are indicated by *t*-test p-values, while the overall differences assessed by the F-test *p*-value from the analysis of covariance model.

**Table 1.** Sample characteristics of the study participants from CHILD, FAMILY, and START cohorts.

|  | **CHILD**<br>**(N=3044)** | **FAMILY**<br>**(N=803)** | **START**<br>**(N=938)** | **ANOVA P-value** |
| --- | --- | --- | --- | --- |
| **Maternal height (cm)** | 165.0 (± 6.9) | 164.3 (± 6.6) | 162.1 (± 6.3) |  |
| Missing | 288 (9.5%) | 6 (0.7%) | 2 (0.2%) | <0.001 |
| **Pre-pregnancy BMI (kg/m<sup>2</sup>)** | 24.3 (± 5.1) | 26.4 (± 6.5) | 23.8 (± 4.4) |  |
| Missing | 1116 (36.7%) | 31 (3.9%) | 2 (0.2%) | <0.001 |
| **Pre-pregnancy weight (kg)** | 66.2 (± 14.9) | 71.4 (± 18.4) | 62.5 (± 11.9) |  |
| Missing | 1078 (35.4%) | 25 (3.1%) | 0 (0%) | <0.001 |
| **Gestational weight gain (kg)** | 15.3 (± 6.3) | 15.3 (± 5.8) | 14.3 (± 7.4) |  |
| Missing | 1059 (34.8%) | 51 (6.4%) | 12 (1.3%) | <0.001 |
| **Mother's Age (years)** | 32.1 (± 4.7) | 32.0 (± 5.2) | 30.2 (± 3.9) |  |
| Missing | 90 (3.0%) | 0 (0%) | 0 (0%) | <0.001 |
| **Parity** | 0.7 (± 0.9) | 0.8 (± 1.0) | 0.8 (± 0.8) |  |
| Missing | 7 (0.2%) | 0 (0%) | 19 (2.0%) | <0.001 |
| **Gestational Diabetes Mellitus** |  |  |  |  |
| Absent | 2942 (96.6%) | 675 (84.1%) | 600 (64.0%) | <0.001 |
| Present | 97 (3.2%) | 128 (15.9%) | 334 (35.6%) |  |
| Missing | 5 (0.2%) | 0 (0%) | 4 (0.4%) |  |
| **Pre-eclampsia** |  |  |  |  |
| Absent | 2944 (96.7%) | 801 (99.8%) | 930 (99.1%) | <0.001 |
| Present | 100 (3.3%) | 2 (0.2%) | 7 (0.7%) |  |
| Missing | 0 (0%) | 0 (0%) | 1 (0.1%) |  |
| **Smoking History** |  |  |  |  |
| Never smoked | 2148 (70.6%) | 485 (60.4%) | 932 (99.4%) | <0.001 |
| Quit before pregnancy | 568 (18.7%) | 112 (13.9%) | 2 (0.2%) |  |
| Quit during pregnancy | 125 (4.1%) | 118 (14.7%) | 2 (0.2%) |  |
| Currently smoking | 96 (3.2%) | 50 (6.2%) | 0 (0%) |  |
| Missing | 107 (3.5%) | 38 (4.7%) | 2 (0.2%) |  |
| **Smoking Exposure (hr/week)** | 1.2 (± 8.7) | 0.4 (± 2.0) | 0.3 (± 2.7) |  |
| Missing | 231 (7.6%) | 11 (1.4%) | 50 (5.3%) | 0.0022 |
| **Years of Education (years)** | 16.6 (± 3.0) | 16.9 (± 3.3) | 16.0 (± 2.5) |  |
| Missing | 131 (4.3%) | 5 (0.6%) | 0 (0%) | <0.001 |
| **Social Disadvantage Index** | 0.5 (± 1.0) | 0.8 (± 1.3) | 1.7 (± 1.4) |  |
| Missing | 498 (16.4%) | 29 (3.6%) | 129 (13.8%) | <0.001 |
| **Plant Based Diet** | -0.5 (± 0.5) | 0.2 (± 0.7) | 1.4 (± 1.1) |  |
| Missing | 274 (9.0%) | 75 (9.3%) | 45 (4.8%) | <0.001 |
| **Health Conscious Diet** | 0.3 (± 1.0) | -0.7 (± 0.7) | -0.4 (± 0.8) |  |
| Missing | 274 (9.0%) | 75 (9.3%) | 45 (4.8%) | <0.001 |
| **Western Diet** | -0.1 (± 0.8) | 1.0 (± 1.2) | -0.6 (± 0.6) |  |
| Missing | 274 (9.0%) | 75 (9.3%) | 45 (4.8%) | <0.001 |
| **Gestational age (weeks)** | 39.7 (± 1.1) | 39.4 (± 1.2) | 39.4 (± 1.1) | <0.001 |
| **Newborn Sex** |  |  |  |  |
| Male | 1591 (52.3%) | 405 (50.4%) | 462 (49.3%) | 0.401 |
| Female | 1453 (47.7%) | 398 (49.6%) | 476 (50.7%) |  |
| **Birth Length (cm)** | 51.6 (± 2.5) | 50.1 (± 2.2) | 51.3 (± 2.4) | |
| Missing | 576 (18.9%) | 26 (3.2%) | 19 (2.0%) | <0.001 |
| **Birth Weight (kg)** | 3.5 (± 0.5) | 3.5 (± 0.5) | 3.3 (± 0.4) | <0.001 |
| **Fat Mass (kg)** | 0.35 (± 0.13) | 0.35 (± 0.13) | 0.36 (± 0.12) | |
| Missing | 1461 (48.0%) | 89 (11.1%) | 113 (12.0%) | 0.006 |
| **Fat Free Mass (kg)** | 3.14 (± 0.34) | 3.13 (± 0.41) | 2.91 (± 0.35) | |
| Missing | 1461 (48.0%) | 89 (11.1%) | 113 (12.0%) | <0.001 |
| **Birth Weight by Length (kg/cm)** | 6.7 (± 0.7) | 6.9 (± 0.8) | 6.4 (± 0.7) | |
| Missing | 576 (18.9%) | 26 (3.2%) | 19 (2.0%) | <0.001 |

There was also a broad pattern of maternal differences, including a higher prevalence of GDM in START (36%) and FAMILY (16%) versus CHILD (3.2%). In fact, all maternal phenotypes were statistically different between cohorts (*p* < 0.001; **Table 1**): South Asian mothers, on average, tended to be younger, shorter in height, had lower BF%, smaller BMI, lower pre-pregnancy weight, had fewer years of education and higher SDI, and mostly followed a plant-based diet compared to both FAMILY and CHILD. Nearly all START mothers were non-smokers (99.4%). In contrast, mothers in FAMILY, on average, had the highest BF%, highest pre-pregnancy BMI, higher pre-pregnancy weight, the most gestational weight gain, the highest rate of smoking, slightly higher SDI than the CHILD cohort, and followed a Western- style diet and were the least healthy in terms of dietary preference.

### Body Compartments Responsible for Difference in BW

When testing specific components of BW, our results revealed significant yet opposing contributions to BW difference (**Figure 1**). While FFM was the primary source of BW difference, contributing to a 0.2kg lower BW in South Asians (*p*<2.2E-16), FM had a significant but weaker effect, contributing to a 0.01kg higher BW in South Asians (*p*=0.006). No difference in weight was observed in early childhood after the 2^nd^ year visit (*p*=0.11; **Table S2**), although a significantly higher skinfold thickness, a measure of FM, at 3 and 5 years were observed in South Asian children (*p* < 0.001; **Table S2**).

### Risk Factors Associated With BW

Across the three studies, a set of 9 maternal characteristics were consistently associated with BW and its components, namely, maternal height, pre-pregnancy weight, pre-pregnancy BMI, weight gain during pregnancy, parity, GDM, maternal glucose AUC, pre-eclampsia, and Western diet (FDR *q*<0.05; Heterogeneity-*p*< 0.05 **Table 2**). However, we found distinct association patterns with different components of BW. For example, pre-eclampsia was only associated with a significant lower BW and FFM. In contrast, GDM and maternal glucose were associated with a significant higher BW and FM. Across the associated maternal characteristics, each 1 SD increase was associated with a birth weight difference ranging roughly between 16–90g (S.E.s=6–7g). All remaining associations involved both the FM and FFM compartments but at slightly different significance and effect sizes (**Tables 2**).

**Table 2.** Association between maternal factors and birth weight.

|  | Difference in distribution<br>(CHILD as reference) |  | Effect Category | Potential causal | Association with BW |  |  | Association with FFM |  |  | Association with FM |  |  |
| --- | --- | --- | --- | --- | --- | --- | --- | --- | --- | --- | --- | --- | --- |
|  | FAMILY | START |  |  | Effect | S.E | Q-value | Effect | S.E | Q-value | Effect | S.E | Q-value |
| Maternal height | - | - | Homogeneous (FM) | YES | 0.09 | 0.006 | 1.31E-46 | 0.08 | 0.006 | 4.11E-43 | 0.01 | 0.002 | 8.06E-05 |
| Pre-pregnancy BMI (kg/m2) | + | - | Homogeneous (BW, FFM) | YES | 0.07 | 0.007 | 5.76E-21 | 0.04 | 0.006 | 6.86E-10 | 0.03 | 0.002 | 3.28E-39 |
| Pre-pregnancy weight (kg) | + | - | Homogeneous (BW, FFM) | YES | 0.09 | 0.007 | 2.29E-42 | 0.06 | 0.005 | 3.19E-27 | 0.03 | 0.002 | 3.44E-44 |
| Gestational weight gain (kg) | N.D | - | Homogeneous (FFM) | YES | 0.05 | 0.007 | 9.04E-15 | 0.04 | 0.006 | 4.74E-13 | 0.01 | 0.002 | 1.76E-05 |
| Mother's Age (years) | N.D | - | Homogeneous | NO | 0.01 | 0.006 | 1.06E-01 | 0.01 | 0.006 | 1.55E-01 | 0.00 | 0.002 | 1.53E-01 |
| Parity | + | + | Homogeneous (BW, FFM) | YES | 0.07 | 0.006 | 1.39E-28 | 0.05 | 0.006 | 3.11E-18 | 0.02 | 0.002 | 3.28E-12 |
| Gestational Diabetes Mellitus | + | + | Homogeneous (FM) | YES | 0.03 | 0.006 | 7.32E-05 | 0.01 | 0.006 | 3.66E-01 | 0.02 | 0.002 | 3.03E-12 |
| Maternal glucose AUC |  |  | Homogeneous (FM) | YES | 0.04 | 0.006 | 1.72E-09 | 0.02 | 0.006 | 1.54E-02 | 0.00 | 0.002 | 4.77E-01 |
| Pre-eclampsia | - | - | Homogeneous | YES | -0.02 | 0.006 | 1.24E-02 | -0.01 | 0.005 | 5.06E-02 | 0.00 | 0.002 | 9.60E-01 |
| Smoking History | + | - | Homogeneous | NO | -0.01 | 0.006 | 3.83E-01 | -0.01 | 0.006 | 4.29E-01 | 0.00 | 0.003 | 1.34E-01 |
| Smoking Exposure (hr/week) | - | - | Homogeneous (FFM) | NO | 0.00 | 0.006 | 8.51E-01 | -0.01 | 0.007 | 1.55E-01 | -0.01 | 0.002 | 6.58E-04 |
| Years of Education | + | - | Heterogeneous | NO | -0.02 | 0.006 | 7.24E-04 | -0.01 | 0.006 | 5.06E-02 | 0.00 | 0.002 | 7.21E-01 |
| Social Disadvantage Index | + | + | Heterogeneous | NO | 0.00 | 0.007 | 5.37E-01 | 0.00 | 0.006 | 7.62E-01 | 0.00 | 0.002 | 2.08E-01 |
| Plant Based Diet | + | + | Homogeneous (FM) | NO | -0.01 | 0.006 | 1.51E-01 | 0.00 | 0.006 | 7.62E-01 | 0.00 | 0.002 | 6.67E-01 |
| Health Conscious Diet | - | - | Homogeneous (BW, FFM) | NO | 0.00 | 0.006 | 5.37E-01 | 0.00 | 0.006 | 8.09E-01 | 0.01 | 0.002 | 2.00E-02 |
| Western Diet | + | - | Homogeneous (BW) | YES | 0.03 | 0.006 | 1.18E-05 | 0.02 | 0.006 | 1.78E-02 | 0.02 | 0.002 | 2.33E-13 |

### Characteristics of PGSs

In general, maternal anthropometric and body composition PGSs (i.e. height, BMI, FM, FFM) performed well, explaining 6–36.3% of the phenotypic variance for the corresponding trait across the genetic subsamples from each cohort (**Table S3**; *p*=8.43×10^-9^–4.39×10^-36^). PGSs related to maternal health conditions (i.e. T2D and fasting glucose) performed better in samples with a higher prevalence of those conditions, such as the START cohort (*p*<1.3×10^-5^). Moreover, maternal education and smoking PGSs were significantly associated with the corresponding outcomes in both CHILD and FAMILY (**Table S3**). However, in the START cohort, the education PGS only showed a weak association with years of education, and the smoking PGS was not significant, likely due to the cohort’s lack of smoking prevalence. The maternal PGSs were strongly associated with their respective traits, confirming their validity as robust genetic instruments for causal inference. Finally, the genome-wide fetal PGS for newborn BW explained 2.1%, 8.1%, and 3.7% of BW variance in the genetic subsamples of CHILD, FAMILY, and START, respectively, confirming the role of fetal genetics on BW.

### Examining Potential Causal Pathways through Polygenic Scores

Using valid genetic instruments, we identified significant causal effects of maternal traits on BW through both the IVW regression and the GMM approach with materially the same results (**Tables S5** and **S6**). Specifically, genetically predicted maternal pre-pregnancy weight and BMI broadly contribute to increased BW, through both FM and FFM (**Table 3**; **Figure S2**), consistent with the direction of the observed associations between maternal traits and BW. For example, we found that a 1 SD increase in mPGS of FFM was associated with an average of 131g (95% CI: 58–204g) increase in BW, with a 103g (95% CI: 45–162) increase in the FFM and 38g (95% CI: 17–60g) increase in FM. In contrast, while GDM and maternal glucose AUC were associated with BW and both compartments, genetically predicted GDM and glucose AUC were only associated with an increase in FM and not FFM. For example, a 1 SD increase in mPGS of T2D was associated with a 60g (95% CI: 20–100g) increase in FM component (*p*=0.008; **Table 3**). In all cases, the significant causal associations showed consistent directions and effect sizes with the corresponding trait associations, except for GDM, which had a much larger effect and SD. Each 1 SD increase in a maternal trait had a direct effect on BW ranging from 36 to 131g (standard errors: 22–46 g). Maternal glucose AUC had a similar estimated effect as GDM, and the results in non-GDM mothers remained virtually the same (**Figure S3**). Using alternative definition of GDM did not change the results (**Figure S4**), with the ethnically appropriate definition (i.e. BiB for SA and IADPSG for European mothers) yielding the strongest effect and the smallest standard error. In the sensitivity analysis, only the fetal genetic effect of height was significantly associated with BW through FFM, supporting a direct fetal genetic effect on growth independent of maternal genetics (**Figure S5**).

**Table 3.** Maternal factors with a causal effect on birth weight and its components.

| Outcome | Maternal Trait | PGS | Pooled Estimate | Pooled Std Error | Pooled CI Lower | Pooled CI Upper | P Value |
| --- | --- | --- | --- | --- | --- | --- | --- |
| Birthweight | BMI | BMI | 0.118 | 0.046 | 0.028 | 0.209 | 0.010 |
| Birthweight | BMI | Fat mass | 0.086 | 0.038 | 0.011 | 0.160 | 0.024 |
| Birthweight | GDM | T2D | 0.256 | 0.105 | 0.049 | 0.462 | 0.015 |
| Birthweight | Glucose AUC | T2D | 0.188 | 0.086 | 0.019 | 0.358 | 0.029 |
| Birthweight | Weight | Fat free mass | 0.131 | 0.037 | 0.058 | 0.204 | 0.00043 |
| Birthweight | Weight | Fat mass | 0.086 | 0.038 | 0.011 | 0.160 | 0.024 |
| Fat Free Mass | BMI | BMI | 0.075 | 0.037 | 0.003 | 0.147 | 0.042 |
| Fat Free Mass | BMI | Fat mass | 0.065 | 0.030 | 0.005 | 0.124 | 0.033 |
| Fat Free Mass | Height | Height | 0.040 | 0.018 | 0.004 | 0.076 | 0.030 |
| Fat Free Mass | Weight | Fat free mass | 0.103 | 0.030 | 0.045 | 0.162 | 0.00051 |
| Fat Free Mass | Weight | Fat mass | 0.065 | 0.030 | 0.005 | 0.124 | 0.033 |
| Fat Mass | BMI | BMI | 0.036 | 0.014 | 0.009 | 0.063 | 0.0095 |
| Fat Mass | GDM | T2D | 0.084 | 0.030 | 0.026 | 0.142 | 0.0043 |
| Fat Mass | Glucose AUC | T2D | 0.058 | 0.022 | 0.015 | 0.101 | 0.0079 |
| Fat Mass | Weight | Fat free mass | 0.038 | 0.011 | 0.017 | 0.060 | 0.00056 |

### Quantifying the Impact of Causal Factors on the BW Gap

Since our genetic analyses confirmed that the effects of the five factors on trait associations (**Table S4**) and direct maternal effects (**Table S5**) to be consistent across studies, we were able to directly pool the data and adjust for the five factors in the regression models. Accordingly, we focused on epidemiological results and whether they corroborate conclusions from the genetic analysis. Accounting for factors with direct maternal effects, the observed BW difference between the SA and White European newborns was reduced to 100g (95% CI: 67–133g; **Figure 2**), 54% of the original difference in BW (217g; 95% CI: 186–249g). The overall reduction in BW was attributable to a combination of increased FM in South Asian and decreased FFM in the White European newborns after adjustment. Specifically, the difference in FM was reduced by 21% (from 233g to 184g), and the difference in FFM was reduced by 56% (from 16 g to 7 g) when adjusting for FFM and FM-specific causal factors. The matched analysis, which included 692 pairs of matched mothers, yielded materially similar results (**Figure 2**).

**Figure 2.**
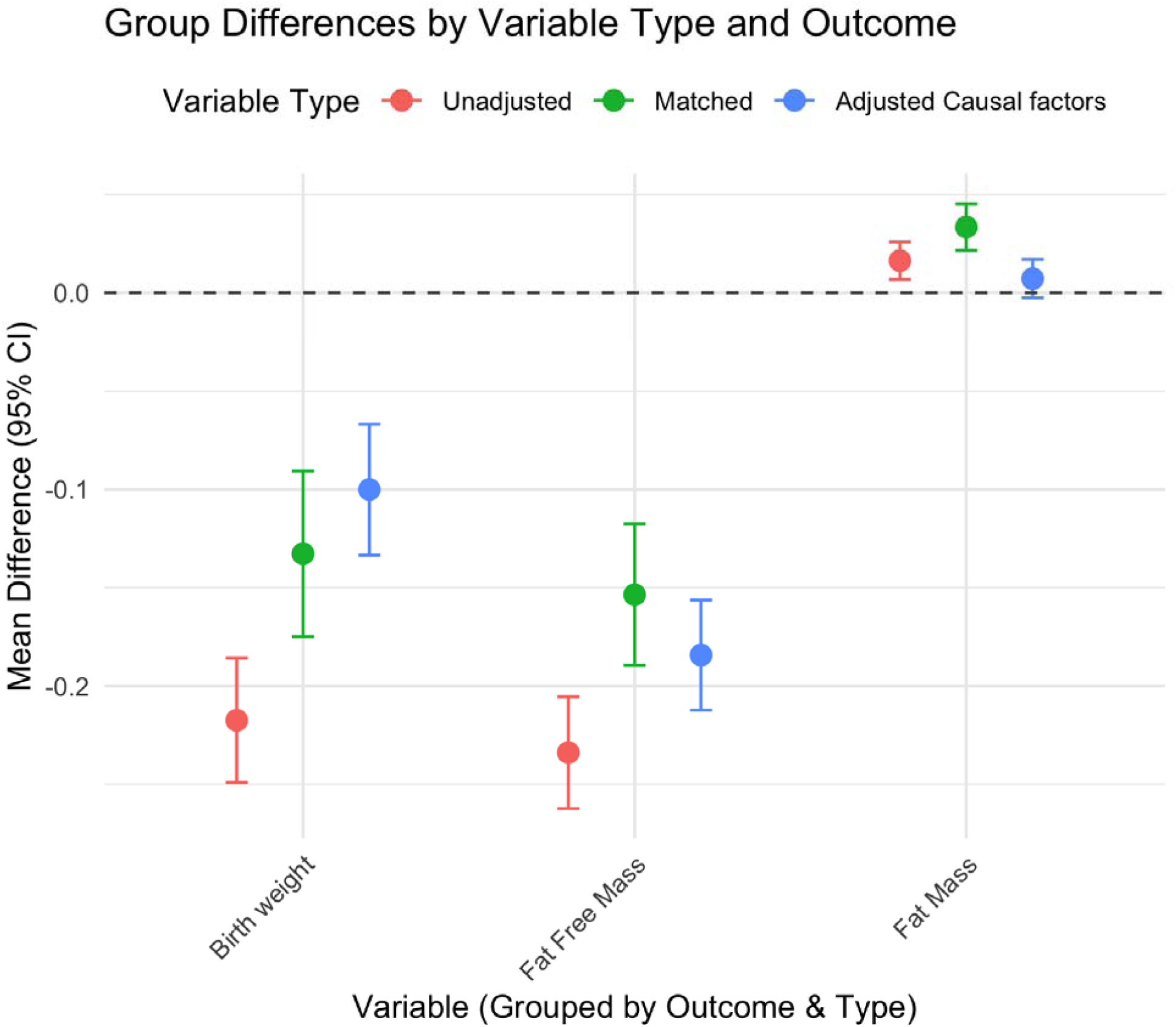
Group differences in birth weight and body composition by adjustment type. Mean differences (with 95% confidence intervals) in birth weight (BW), fat-free mass (FFM), and fat mass (FM) between South Asian and White European newborns are shown under three models: unadjusted (red), matched analysis (green), and adjusted regression model including identified causal maternal factors (blue). Negative values indicate lower values in South Asian newborns relative to White European newborns. After adjustment, the mean difference in BW was substantially reduced, through a reduction in the negative FFM difference and in the positive FM difference between South Asian and White European newborns.

## Discussion

In this study, we confirmed the difference in BW between South Asian and predominantly White European newborns was driven by a lower FFM and a slightly higher FM in South Asian newborns. The maternal causal drivers that significantly contributed to each compartment of BW difference included maternal weight, height and glucose during pregnancy.

Our findings have three implications: 1) The lower BW in South Asians is primarily due to a lower muscle mass and presumably bone mass, which are critical components of FFM. Given the ancestral origin of the low FFM phenotype^60^, it is unsurprising that the low BW persists in South Asians in India^6,8^ and also South Asian offspring generations after immigration^22^. A low FFM has been suggested as the culprit for the “obesity paradox”^61^, whereby an association between a lower BMI and all-cause mortality was observed but possibly due to lower FFM rather than lower overall weight^62^. 2) Different maternal factors causally influence BW and its components, with distinct pathways driving fetal growth. Further, these maternal factors are often correlated and should not be considered in isolation. For example, when stratified by maternal BMI deciles alone, we observed instances where there was no ethnic difference in overall BW, but significantly higher FM and significantly lower FFM in South Asian newborns (**Figure S5**). This suggests that ethnic differences in neonatal body composition exist even when maternal BMI is controlled for. More importantly, these findings emphasize the need to move beyond BW as a singular measure of neonatal health and consider the composition of BW, which may have metabolic consequences. 3) A higher prevalence of GDM in South Asian mothers may contribute to their infants having greater neonatal adiposity despite lower overall BW. Over time, this lower FFM and higher adiposity%^24,63^ are preserved in South Asian children, even as the overall weight differences diminish with age. This, in turn, might predispose them to an elevated risk of metabolic disorders later in life—potentially perpetuating the cycle of maternal GDM risk in future generations^26,27,64^.

The observation that body composition, as well as GDM and maternal glucose drive FM accumulation is particularly relevant for understanding ethnic differences in neonatal body composition. South Asian mothers were more likely to have a lower body weight, lower BMI and higher rate of GDM, contributing to an overall lower BW and higher FM. Our findings point to potential interventions targeting both body composition (weight, height, adiposity) and maternal glucose regulation could be particularly beneficial for reducing neonatal adiposity and its long-term consequences. However, we did not find any actionable mechanisms that can increase FFM without increasing FM in the present study. While maternal height was previously identified as an important determinant of BW, our result suggests that its role is largely restricted to FFM, with minimal overall impact after accounting for gestational age and newborn sex. Similarly, education and diet are strongly associated with BW and its components, but a direct maternal influence of these traits on BW was not supported.

Strengths of this study include its large sample size, high quality maternal phenotypes, genetic causal inference approach, and comparison across three well-characterized Canadian birth cohorts, which enhance the robustness of the findings. South Asians already represent the largest visible minority group in Canada and the largest non-White ethnic group in the country^65^.

Similarly, sizable and expanding South Asian communities are found in the United Kingdom, United States, Australia, and parts of the Caribbean and Africa. Our findings are directly relevant to healthcare systems in Canada and globally, where culturally and ancestrally informed strategies will be essential for reducing long-standing health disparities.

However, limitations and potential future directions should also be considered. First, although we accounted for key maternal factors such as adiposity, diet, and clinical variables, other important factors like maternal stress^66^ and air pollution^67^ that were not captured in all cohorts, could influence fetal growth. Second, genetic instruments may not fully capture all aspects of maternal traits, and differences in nutritional status and healthcare access could influence BW and body composition in ways not fully accounted for. Third, comparative studies in adults, preferably as a follow up in the same birth cohorts, are needed to replicate the associations between genetic factors and the observed FFM and adiposity in South Asian and White Europeans. Investigating the long-term cardiometabolic consequences of these early differences in body composition is crucial to provide a more comprehensive understanding of how to mitigate health disparities.

In conclusion, this study contributes to a growing body of evidence emphasizing the importance of metabolic health in pregnancy and the need to move beyond BW as a singular measure of neonate body composition. Our findings provide a strong rationale for integrating maternal glucose regulation and weight management into perinatal care strategies to improve long-term health outcomes for both mothers and their offspring. Given the disproportionately high prevalence of GDM and relative neonatal adiposity among South Asians, targeted interventions in this population may be essential for breaking the cycle of intergenerational metabolic risk.

## Contributors

Wei Q. Deng contributed to conceptualization, investigation, methodology, formal analysis, visualization, writing of the original draft, and review and editing of the manuscript. Marie Pigeyre contributed to investigation and review and editing of the manuscript. Sandi M. Azab contributed to investigation and manuscript review and editing. Amel Lamri contributed to data curation, investigation, and manuscript review and editing. Russell J. de Souza contributed to investigation and review and editing of the manuscript. Natalie Williams contributed to data curation and project administration. Kyla Belisario contributed to investigation and manuscript review and editing. Nathan Cawte contributed to data curation, review and editing of the manuscript. Dipika Desai contributed to project administration and data curation. Katherine M. Morrison, Stephanie A. Atkinson, Koon K. Teo, Theo J. Moraes, Padmaja Subbarao, Stuart E. Turvey, Piush Mandhane, and Elinor Simons contributed to data curation, investigation, and manuscript review and editing. Guillaume Paré contributed to data curation, methodology, supervision, and manuscript review and editing. Sonia S. Anand contributed to conceptualization, methodology, supervision, funding acquisition, data curation, and review and editing of the manuscript. All authors reviewed and approved the final version of the manuscript. Wei Q. Deng had full access to all the data in the study and verify the integrity and accuracy of the data analysis.

## Data Availability

The PGS data used to derive the PGS are available from the PGS catalog with reference number shown in Table S3. Data collected for this study, including individual participant data and data dictionary defining each field in the dataset, are available to researchers who provide a proposal detailing intended analyses, pending access approval by the NutriGen Alliance Team. CHILD study data can be separately requested from the CHILDdb database. Access can be initiated through https://childstudy.ca/childdb/.

## Acknowledgments

We express our sincere gratitude to all the participating families and the START, FAMILY and CHILD study teams, including interviewers, nurses, computer and laboratory technicians, clerical workers, research scientists, volunteers, managers, and receptionists. We thank the South Asian Birth Cohort (START) study, Family Atherosclerosis Monitoring in Early Life (FAMILY) study, and CHILD Cohort Study (CHILD) participant families for their dedication and commitment to advancing health research. CHILD was initially funded by the Canadian Institutes of Health Research, Debbie and Don Morrison and the Allergy, Genes, and Environment Network of Centres of Excellence (AllerGen NCE), Women’s and Children’s Health Research Institute provided core support for establishing the CHILD Study. Visit CHILD at childstudy.ca. The Family Atherosclerosis Monitoring in Early Life (FAMILY) study was supported by the Canadian Institutes of Health Research, the Population Health Research Institute, and the McMaster Children’s Hospital Foundation. The South Asian Birth Cohort (START) study was supported by the Indian Council of Medical Research and in Canada by the Canadian Institutes of Health Research (grant INC-109205), and the Heart and Stroke Foundation (grant NA7283).

We would like to acknowledge the Genetic and Molecular Epidemiology Laboratory (GMEL), an associate of Hamilton Health Sciences and McMaster University, for their indispensable contributions to this work. The technical staff of GMEL conducted all epigenetic profiling, including sample processing and other technical operations.

We thank the members of the Nutrigen Alliance for providing the data: Sonia S. Anand; Stephanie A. Atkinson; Meghan Azad; Allan B. Becker; Jeffrey Brook; Judah A Denburg; Dipika Desai; Russell J. de Souza; Milan K. Gupta; Michael Kobor; Diana L. Lefebvre; Wendy Lou; Piushkumar J. Mandhane; Sarah McDonald; Andrew Mente; David Meyre; Theo J. Moraes; Katherine M. Morrison; Guillaume Paré; Malcolm R. Sears; Padmaja Subbarao; Koon K. Teo; Stuart E. Turvey; Julie Wilson; Salim Yusuf; Gita Wahi; Michael A. Zulyniak.

This research was supported by the Canadian Institutes of Health Research DOHaD Team Grant (MWG-146332). Dr. Anand is supported by a Tier 1 Canada Research Chair in Ethnicity and CVD and Heart, Stroke Foundation Chair in Population Health, a grant from the Canadian Partnership Against Cancer, Heart and Stroke Foundation of Canada, and Canadian Institutes of Health Research. Dr. Subbarao is supported by a Tier 1 Canada Research Chair in Pediatric Asthma and Lung Health. Dr. Turvey is supported by a Tier 1 Canada Research Chair in Pediatric Precision Health.

Generative AI (ChatGPT, OpenAI, GPT-4, May 2025 version) was used to assist in improving the clarity, readability, and grammar of the manuscript text. Prompts included requests such as: “Rephrase this sentence for clarity” and “Format author contributions using CRediT taxonomy.”

Other generative AI (Claude 3.7 Sonnet) was used to condense portions of the text (abstract and Introduction) to meet word count requirements while preserving the key findings, methods, and conclusions. The AI helped restructure and prioritize content without altering the scientific substance or accuracy of the research findings. These models were not used to generate any scientific content, conduct data analysis, interpret results, or draw conclusions. All AI-assisted output was reviewed and edited by the authors for accuracy and appropriateness. The authors maintained full responsibility for the scientific integrity and content of the manuscript.

## Data sharing statement

The PGS data used to derive the PGS are available from the PGS catalog with reference number shown in **Table S3**. Data collected for this study, including individual participant data and data dictionary defining each field in the dataset, are available to researchers who provide a proposal detailing intended analyses, pending access approval by the NutriGen Alliance Team. CHILD study data can be separately requested from the CHILDdb database. Access can be initiated through https://childstudy.ca/childdb/.

## Abbreviations

ANOVA: Analysis of Variance
BF%: Body Fat Percentage
BW: Birth Weight
CHILD: Canadian Healthy Infant Longitudinal Development Study
FAMILY: Family Atherosclerosis Monitoring in Early Life Study
FFM: Fat-Free Mass
FM: Fat Mass
fPGS: Fetal Polygenic Score
GDM: Gestational Diabetes Mellitus
GWAS: Genome-Wide Association Study
mPGS: Maternal Polygenic Score
OGTT: Oral Glucose Tolerance Test
PGS: Polygenic Score
SDI: Social Disadvantage Index
START: South Asian Birth Cohort Study
T2D: Type 2 Diabetes

---

**Figure S1.**
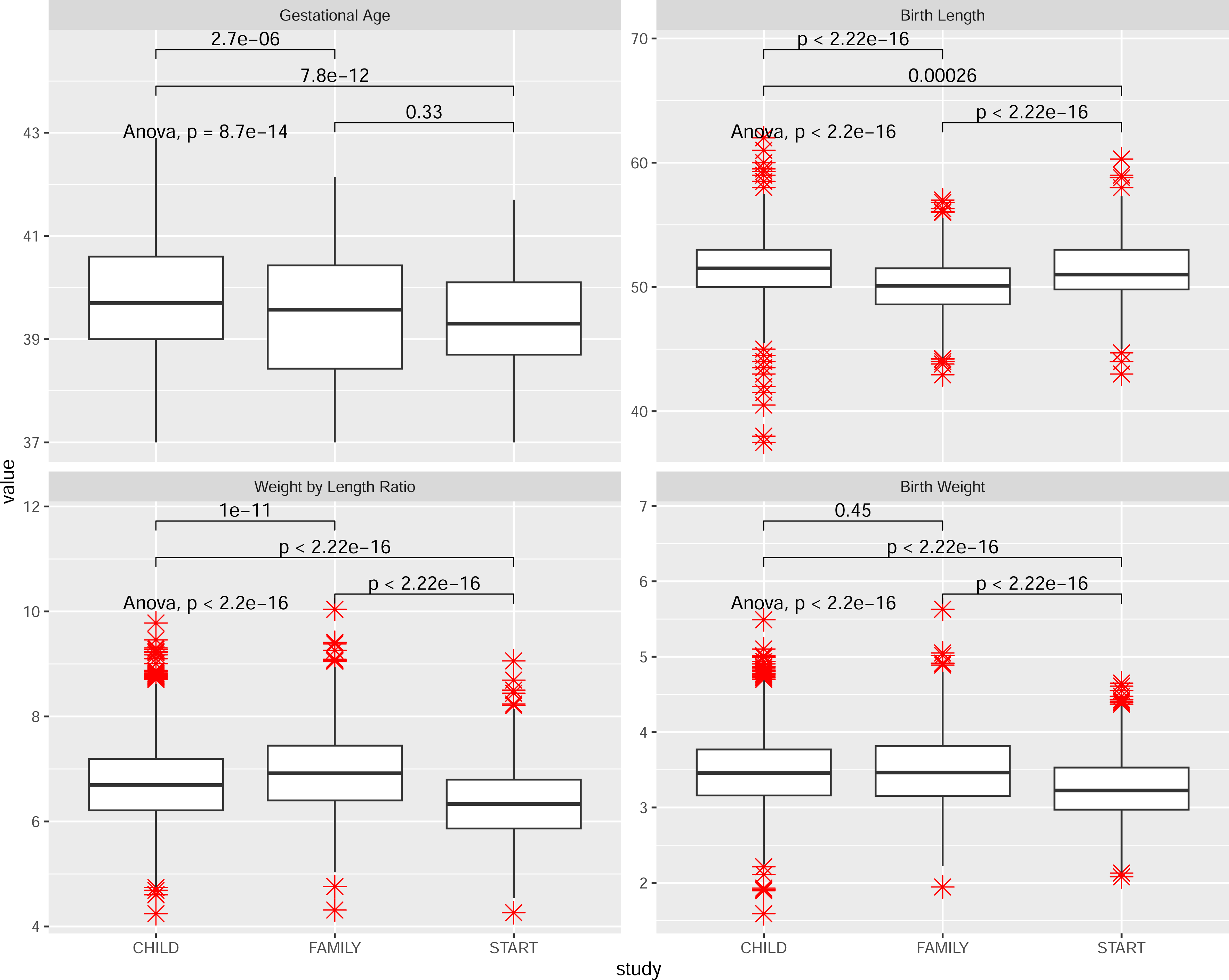
Stratified boxplots of gestational age, birth length, birth weight by length, and birth weight across three studies: CHILD, FAMILY, and START. Each boxplot illustrates the distribution of outcomes within each study cohort. Pairwise comparisons between studies are indicated by *t*-test p-values, while the overall differences assessed by the F-test *p*-value from the analysis of covariance model.

**Figure S2.**
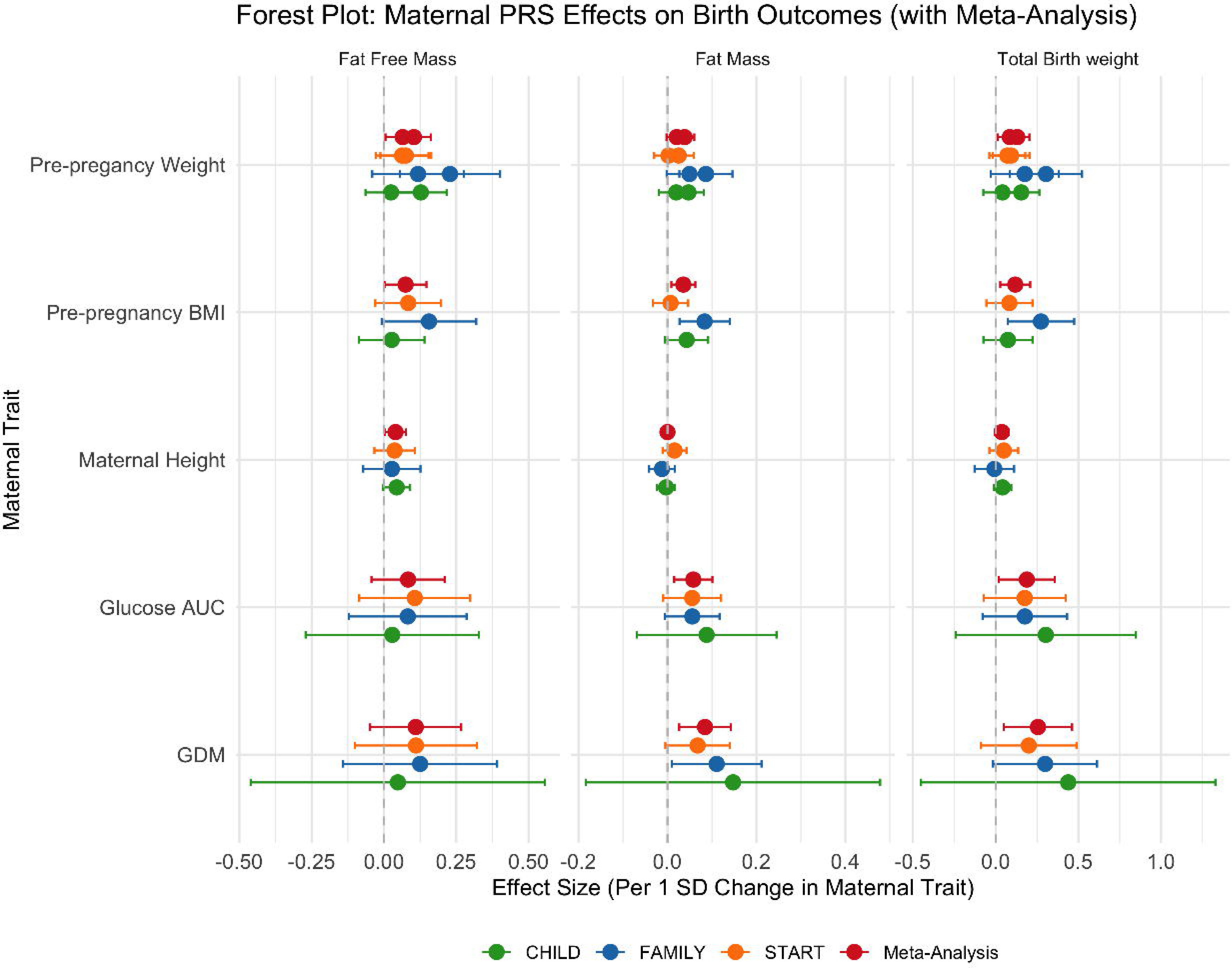
Forest plots of meta-analysis of direct maternal influence on birth weight, fat-free mass, and fat mass. This figure illustrates the direct effect of maternal factors via polygenic risk scores (mPGS) on birth weight components, including fat-free mass (FFM), fat mass (FM), and total birth weight (BW), across three independent cohorts (CHILD, FAMILY, and START) and a meta-analysis that aggregates the findings. The x-axis represents the effect size, quantified as the change in birth outcomes per 1 standard deviation (SD) increase in mPGS, while the y-axis lists the maternal traits analyzed: pre-pregnancy weight, pre-pregnancy BMI, maternal height, maternal blood glucose area under the curve, and gestational diabetes mellitus. Each dataset is represented by a different color: green for CHILD, blue for FAMILY, orange for START, and red for the meta-analysis, which provides a combined estimate of effect sizes across the three cohorts. Error bars indicate 95% confidence intervals (CIs), with wider intervals reflecting greater variability or uncertainty in effect estimates.

**Figure S3.**
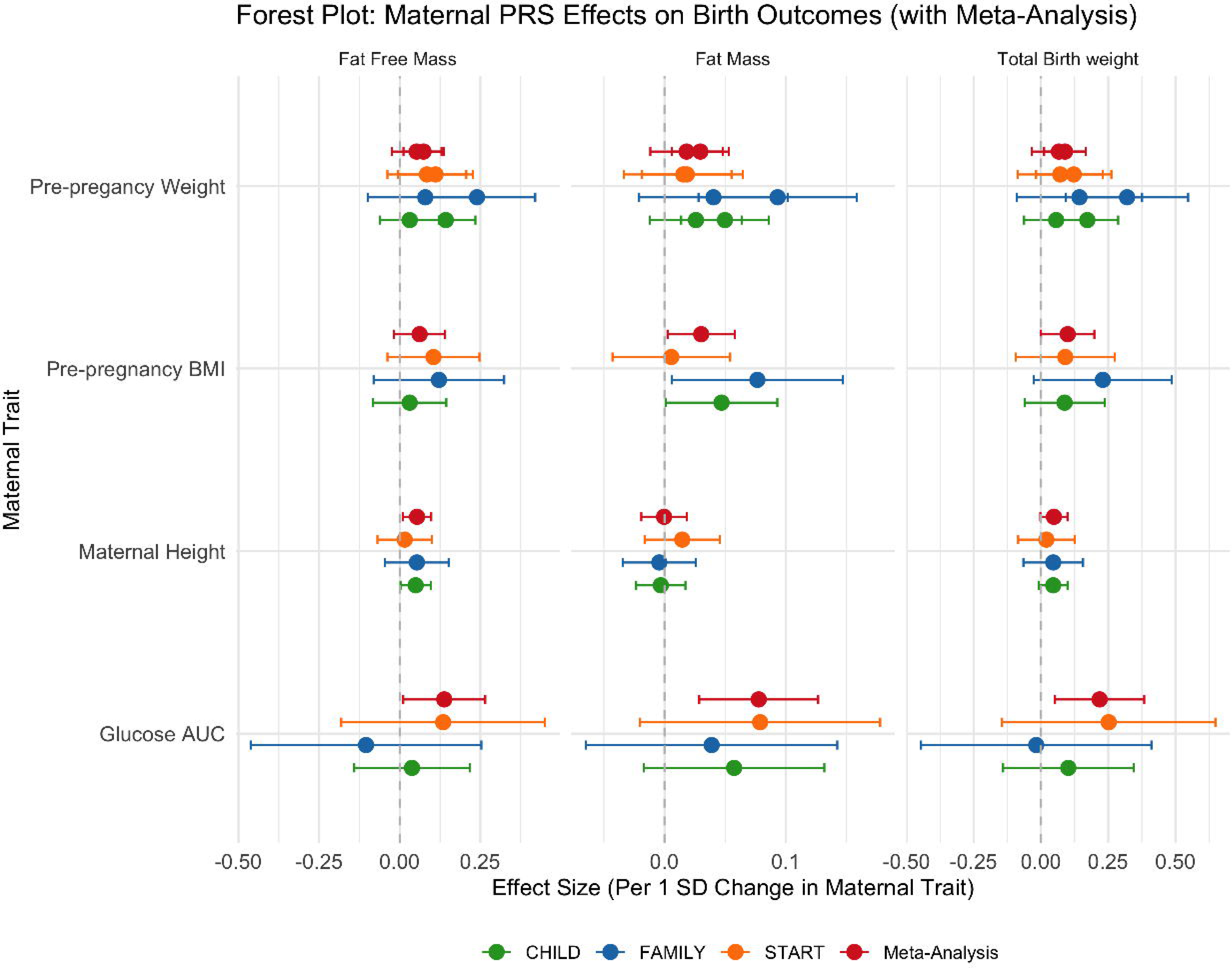
Forest plots of meta-analysis of direct maternal influence on birth weight, fat-free mass, and fat mass in non-GDM mothers. This figure illustrates the direct effect of maternal factors via polygenic risk scores (mPGS) on birth weight components, including fat-free mass (FFM), fat mass (FM), and total birth weight (BW) restricted to non-GDM mothers. The x-axis represents the effect size, quantified as the change in birth outcomes per 1 standard deviation (SD) increase in mPGS, while the y-axis lists the maternal traits analyzed: pre-pregnancy weight, pre-pregnancy BMI, maternal height, maternal blood glucose area under the curve, and gestational diabetes mellitus. Each dataset is represented by a different color: green for CHILD, blue for FAMILY, orange for START, and red for the meta-analysis, which provides a combined estimate of effect sizes across the three cohorts. Error bars indicate 95% confidence intervals (CIs), with wider intervals reflecting greater variability or uncertainty in effect estimates.

**Figure S4.**
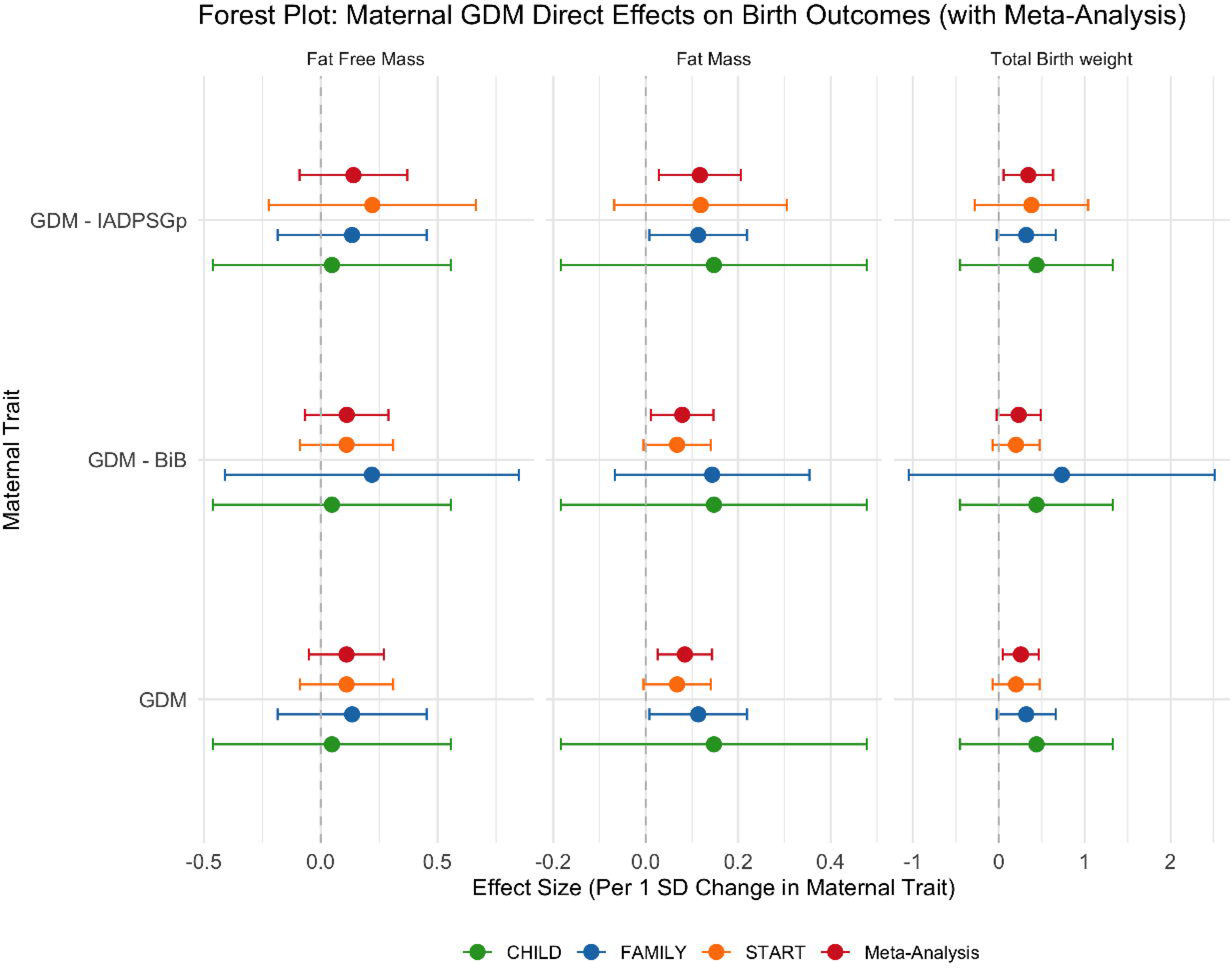
Forest plots of meta-analysis of direct maternal influence on birth weight, fat-free mass, and fat mass using alternative GDM definitions. This figure illustrates the direct effect of maternal factors via polygenic risk scores (mPGS) on birth weight components, including fat-free mass (FFM), fat mass (FM), and total birth weight (BW). The x-axis represents the effect size, quantified as the change in birth outcomes per 1 standard deviation (SD) increase in mPGS, while the y-axis lists the different definitions of GDM analyzed: both FAMILY and START using the IADPSG definition, both FAMILY and START using the Born in Bradford definition, the ethnic appropriate definition in FAMILY and START. Each dataset is represented by a different color: green for CHILD, blue for FAMILY, orange for START, and red for the meta-analysis, which provides a combined estimate of effect sizes across the three cohorts. Error bars indicate 95% confidence intervals (CIs), with wider intervals reflecting greater variability or uncertainty in effect estimates.

**Figure S5.**
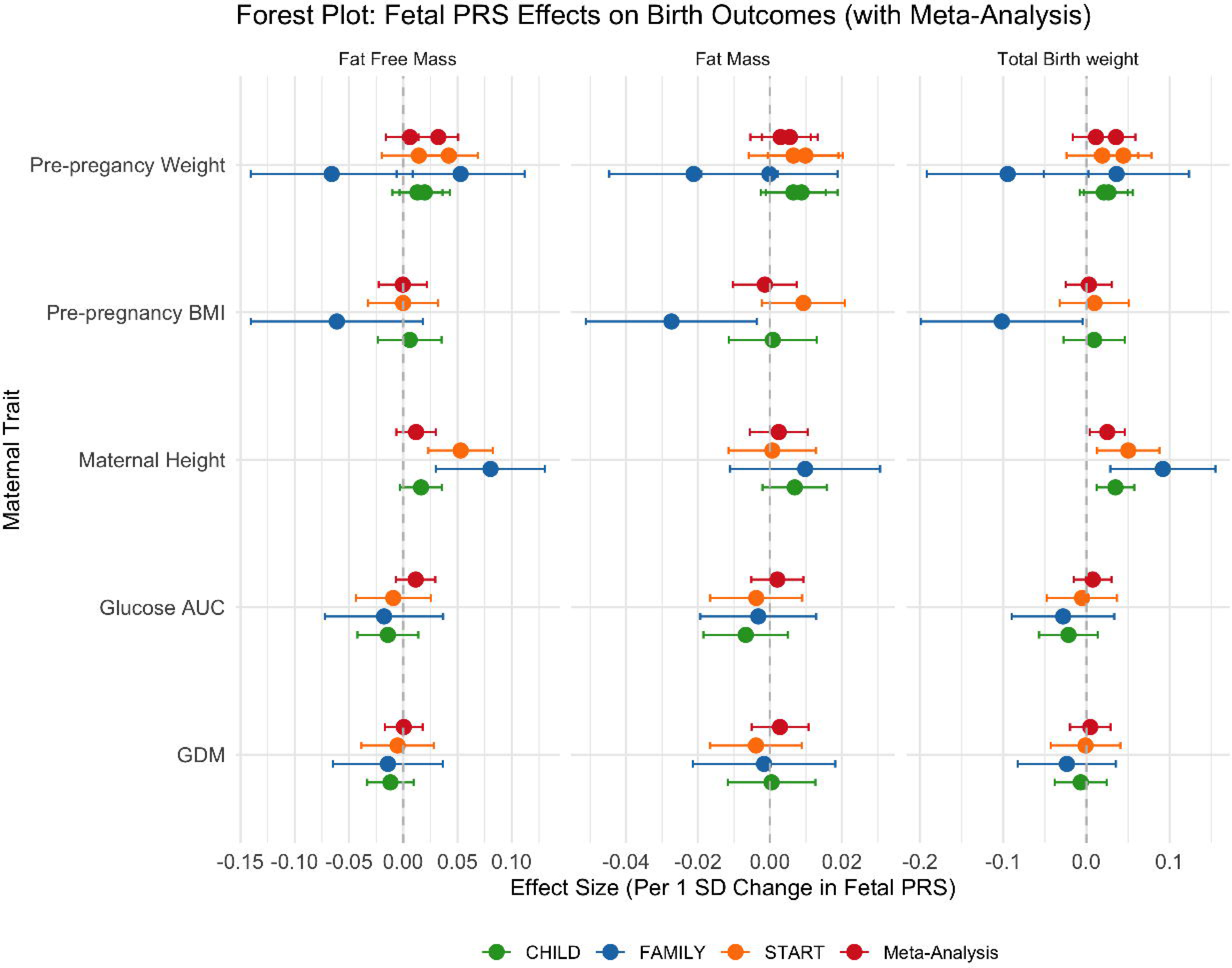
Forest plots of meta-analysis of direct fetal influence on birth weight, fat-free mass, and fat mass. This figure illustrates the direct effects of fetal polygenic risk scores (fPGS) on birth weight components, including fat-free mass (FFM), fat mass (FM), and total birth weight (BW), across three independent cohorts (CHILD, FAMILY, and START) and a meta-analysis that aggregates the findings. The x-axis represents the effect size, quantified as the change in birth outcomes per 1 standard deviation (SD) increase in fPGS, while the y-axis lists the maternal traits analyzed: pre-pregnancy weight, pre-pregnancy BMI, maternal height, maternal blood glucose area under the curve, and gestational diabetes mellitus. Each dataset is represented by a different color: green for CHILD, blue for FAMILY, orange for START, and red for the meta-analysis, which provides a combined estimate of effect sizes across the three cohorts. Error bars indicate 95% confidence intervals (CIs), with wider intervals reflecting greater variability or uncertainty in effect estimates.

**Figure S6.**
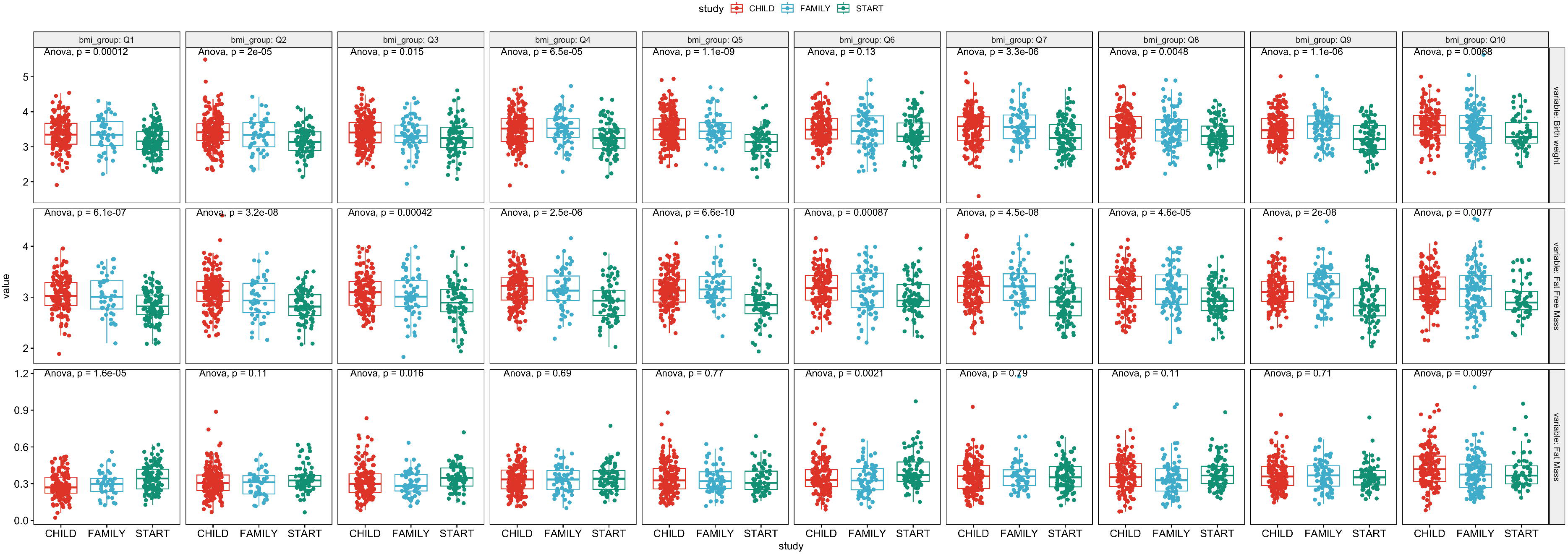
Birth weight and body composition differences across maternal BMI deciles, stratified by study cohort. Boxplots display birthweight (top row), fat-free mass (FFM; middle row), and fat mass (FM; bottom row) by maternal BMI decile (Q1–Q10) across three birth cohorts: CHILD (red), FAMILY (blue), and START (green). Each column represents one maternal BMI decile group. Within each decile, ethnic and cohort differences are compared using ANOVA, with corresponding F-test *p*-values shown above each panel. Notably, in the 6th BMI decile, birthweight differences between cohorts are minimal or non-significant, while significant differences in FM and/or FFM remain, indicating discordant infant body composition profiles despite similar overall birth weight.

