## Supplementary Methods for "Maternal Profiles Account for Birth Weight Differences Across Ethnicities: Results from Three Canadian Birth Cohorts"

Supplementary Materials for “Birth weight disparity”

### A) Phenotype curation and processing

Specifically, newborn length was measured using the O’LEARLY length board. Waist and hip circumferences, and height at later visits, were measured using an OHAUS non-stretchable tape with an attached spring balance. Triceps and subscapular SFT were measured three times using the HOLTAIN callipers in START (a precision of 0.2 mm) and the LANGE callipers in FAMILY (a precision of 0.5 mm). The final reported SFT was the average of three measurements and rounded to the nearest mm.

#### Slaughter’s body fat % estimation in newborn and young children

Body fat percentage (BF%) at birth and follow-up visits was estimated using the Slaughter skinfold-thickness equations (Slaughter et al., 1988) without calf skinfold:

If subcutaneous triceps skinfold < 35mm, then

Boys % Body Fat = 1.21* (Triceps skinfold) -0.008* (Triceps skinfold^2) -1.7

Girls % Body Fat = 1.33* (Triceps skinfold ) -0.013* (Triceps skinfold^2)-2.5;

else

Boys % Body Fat = 0.783* (Triceps skinfold) +1.6

Girls % Body Fat = 0.546* (Triceps skinfold ) + 9.7.

#### Predicted body fat % estimation in newborn from CHILD

Since only FAMILY and START had measures of newborn skinfold thickness for estimating BF%, we sought to predict BF% in newborns from CHILD using models trained in FAMILY, and then compared to an alternative method to estimate newborn BF% that does not require skinfold thickness measurements (Lingwood et al., 2012).

The FAMILY data containing all phenotype data relevant for prediction (n = 759): gestational age, newborn sex, newborn length, birth weight, newborn Ponderal Index (PI), and maternal predictors, including pre-pregnancy BMI, GDM status, maternal exercise during pregnancy, pregnancy weight gain. These variables are available in all cohorts for external validation. The FAMILY data were first split 80-20 for training and testing, and a five-fold cross-validation was done in the training data to select the best model. We first pre-processed data using KNN missing data imputation and then fed the training data to 11 prediction algorithms using R package “caret”: earth for Multivariate Adaptive Regression Splines (MARS), bagEarth for Bagging (Bootstrap Aggregating) of MARS, brnn for Bayesian Regularized Neural Networks, avNNet for Model Averaged Neural Networks, monmlp for Monotone Multi-Layer Perceptrons, mlp for standard Multi-Layer Perceptrons, mlpWeightDecay for Multi-Layer Perceptrons with Weight Decay Regularization, bridge for Bayesian Ridge Regression, enet for Elastic Net Regression, glmnet for Generalized Linear Models with Elastic Net Regularization, and svmLinear3 for Support Vector Machines with a Linear Kernel. The out-of-sample performance was assessed using adjusted R2 in the testing data. The Bayesian Regularized NN model gave the best performance, explaining 42.6% of the variation of birth BF% in the hold-out testing data. The remaining methods gave similar performance with variance explained between 23–42%. The final model is available at <https://github.com/WeiAkaneDeng/EpigeneticResearch>, and was used to derive the BF% in CHILD and START. The predicted BF% in START explained 26% of the estimated BF% by the Slaughter’s equation.

On the other hand, the Lingwood equation (Lingwood et al., 2012) involves only birth weight, gender and birth length:

FFM = 0.057 + 0.646 * weight (kg) - 0.089 * gender (1 = male; 2 = female) + 0.009 * length (cm)

FM = weight - FFM

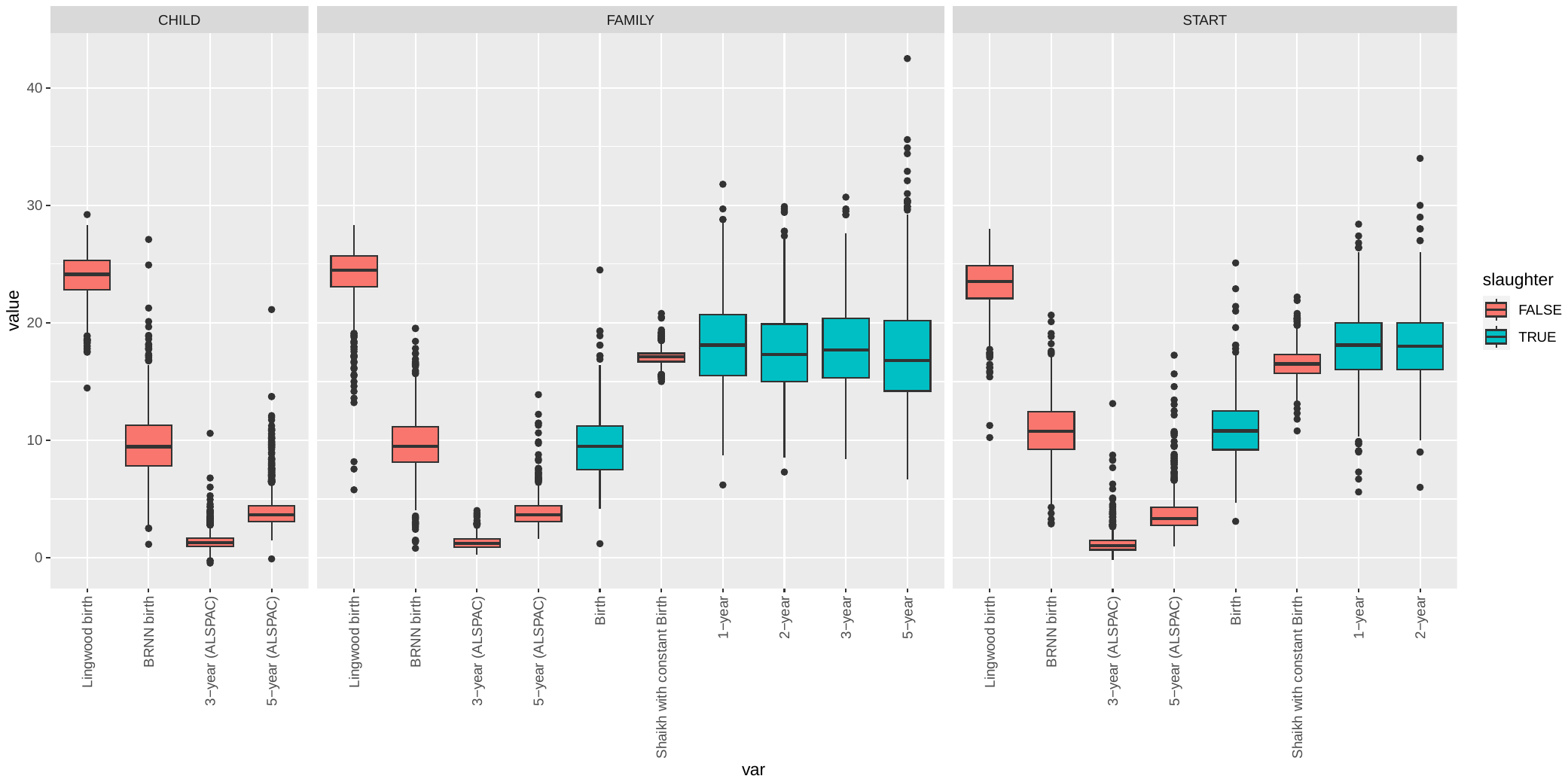

The boxplot above shows the distribution of newborn BF% estimated using different methods across the cohorts.

### B) Genome-wide genotype data

#### Data quality control

Samples from the three cohorts were genotyped using a combination of Illumina’s HumanCoreExome-12 (v1.1), HumanCoreExome-24 (v1.0) and InfiniumCoreExome-24(v1.1) (Illumina Inc, San Diego, CA, USA) all at . Data was cleaned using the following quality control (QC) steps: Participants with low genotyping success rate (<5%), sex discrepancies, high heterozygosity rates, duplicated samples or mother-offspring duos with high numbers of reported Mendel errors were removed. The genetic ancestry of participating was ascertained through mapping genetic principal components using the 1000 Genomes Project (Auton et al., 2015) as reference populations, and samples displaying a discrepancy between self-reported and genetic ancestry were removed. Where first degree relatives were detected, only one arm of the family was kept. Variants with low genotyping success rate (>5%) were excluded and variants with a MAF < 1% and/or HWE ≤ 5.7 × 10-7 were removed prior to imputation. START data was phased using SHAPEIT2 (Delaneau et al., 2011) and imputed using IMPUTE2 (Howie et al., 2009) with the 1000 Genomes phase 3 data as a reference panel (Auton et al., 2015). Samples of European ancestry from CHILD and FAMILY were pre-phased using the Eagle algorithm (Loh et al., 2016), and imputed using Minimac4 (Fuchsberger et al., 2015) via TOPMed’s imputation server (Das et al., 2016) and using the TOPMed r2 release as the reference (Taliun et al., 2021). Variants with a low imputation quality were filtered out before analysis (info score < 0.7 in START and Rsq < 0.3 in CHILD and FAMILY’s European samples).

The following summarizes the data prior to imputation and the imputation procedures:

|  | START | CHILD | FAMILY |
| --- | --- | --- | --- |
| Genotyping arrays | HumanCoreExome-12, HumanCoreExome-24, InfiniumCoreExome-24 | | |
| Ethnicity | South Asian | Multi-ethnic | Multi-ethnic |
| N samples Before QC | 1,506 | 5,229 | 1358 |
| N SNPs before QC | 557,006 | 557,006 | 542,541 |
| N samples that passed QC | 1,449 | 3,651 samples of European ancestry | 1,095 samples of European ancestry |
| Phasing and imputation, softwares and reference panel | SHAPEIT2, IMPUTE2, 1000 Genomes phase 3 reference panel | Eagle, (Minimac4 (v1.6.6) TOPMed reference panel (all populations) | Eagle, (Minimac4 (v1.6.6) TOPMed reference panel (all populations) |

#### Polygenic risk score construction

In particular, we examined 2 reported PRSs of BW that had been separately validated in white European and South Asian populations: one based on genome-wide significant SNPs (Nongmaithem et al., 2022), and the other based on a well-established PRS method “LDpred2” (Privé et al., 2020). The more predictive fetal BW PRS across the cohorts was included in the final analysis. Considering the role of maternal stature and other pregnancy-related complications on BW, we also included maternal PRSs of BW, height, lean BW, fat BW, BMI, T2D, GDM, pre-eclampsia, and glucose level. Finally, we included PRSs of education and smoking as the genetic component of maternal behavioral phenotypes that could also be relevant to BW. All PRSs, except for GDM, were curated from the Polygenic Score (PGS) Catalog (Lambert et al., 2024), selecting the most predictive out of all available PRS for each target phenotype. The best PRS was selected based on performance measures including area under the operating characteristic curve or partial R^2^, the difference in coefficient of correlation R^2^ between the full model and model with the first 20 genetic principal components (PCs) covariates, in out-of-sample testing.

In addition to the genetic QCs, we performed additional QC steps to ensure the reference alleles matched between each discovery GWAS and the target maternal/fetal genotype data from each cohort. PRSs for all traits. For in-sample performance, we calculated the adjusted R^2^ and the *p*-value associated with PRS in a linear regression model with the exact phenotype as the outcome, adjusting for all covariates. We additionally calculated the area under the receiver operating characteristic curve (ROC) for binary outcomes. A nominal *p*-value threshold (*p* < 0.05) was used to determine whether a PRS demonstrated any statistical evidence for association with the target phenotype. Note that for many of the disease outcomes, we expect the association to be close to null due to the low case counts (e.g. pre-eclampsia and GDM in the European cohorts). Finally, fasting blood glucose was not available in any of the cohorts, thus 2-hour post oral glucose tolerance test (OGTT) glucose was used for in-sample performance evaluation in FAMILY and START, while any report of GDM was used as the evaluation phenotype in CHILD.
